# Effects of a personalized or generic three-dimensional tumoral kidney model on patient experience and caregiver-patient interactions, before and after partial nephrectomy, a randomized trial (Rein 3D Print Personalize - UroCCR 114)

**DOI:** 10.1101/2025.04.11.25325651

**Authors:** Maxime Pattou, Sarah Masanet, Marthe-Aline Jutand, Hélène Hoarau, Joffrey Sarrazin, Alice Pitout, Laura Richert, Hugo Larribere, Federico Rubat Baleuri, Manon Jaffredo, Solène Ricard, Matthieu Faessel, Jocelyn Sabatier, Jean-Christophe Bernhard, Gaëlle Margue

## Abstract

**Introduction:** Personalized 3D-printed kidney models could serve as valuable educational tools for patients undergoing robot-assisted partial nephrectomy. These models facilitate patients’ understanding of their pathology, surgical procedure, and anatomy. However, the costs associated with ‘personalized’ printing remain a barrier to their use. This study aims to thoroughly investigate the benefits of using a personalized 3D-printed kidney model as opposed to a generic 3D-printed kidney model as an educational tool for patients and as a communication tool for healthcare professionals.

**Methods and analysis:** In this prospective, single center study, 60 patients undergoing RAPN will be randomized to receive information based on their personalized 3D-printed tumoral kidney model or a generic 3D-printed tumoral kidney model. These models will accompany patients throughout their care pathway, from pre-operative consultations to the post-operative visit. The impact of these models on the management approaches of various healthcare professionals will also be examined. The data will be collected and analyzed using a mixed method, combining interviews (with patients and caregivers), observations during the presentation of the models and questionnaires (understanding of their pathology and the surgical procedure, Health-literacy, satisfaction).

**Discussion:** Three dimensional kidney models have the potential to play a central role in the preoperative information process and serve as an effective educational tool during the patient’s social interactions with relatives and healthcare professionals. This study will evaluate the potential advantages of personalized 3D models of tumoral kidneys compared to their generic counterparts.

**Trial Registration:** The PERSONALIZE study was registered on ClinicalTrials.gov (NCT06035211) on the 5^th^ of September 2023.

## INTRODUCTION

Educational mediation tools to reduce anxiety and improve patient comprehension remains underexploited in modern surgical practice. A pilot study from 2015 demonstrated the benefits of personalized 3D-printed kidney models as educational tools during pre-operative consultations for patients undergoing robot-assisted partial nephrectomy (RAPN) [1] for kidney cancer. This approach enhanced patients’ understanding of their pathology and the surgical procedure, resulting in higher satisfaction levels [2]. In the context of personalized medicine, it is crucial to acknowledge the heterogeneity of patients’ cultural and social backgrounds to optimize the doctor-patient relationship [3]. Patients with lower literacy levels often exhibit more preoperative anxiety [4,5] and poorer post-operative recovery outcomes [6]. Numerous studies have aimed to enhance the health literacy of cancer patients and improve doctor-patient communication [7–9].

Patients diagnosed with renal tumors interact with multiple healthcare professionals, and the nature of their interactions can be influenced by various factors. These include patient-related characteristics (such as clinical setting, context, age, gender, occupation, and literacy levels) as well as healthcare professional-related perceptions, like viewing patients as active participants in their care, which can alter relations and expectations [10].

New digital technologies improve patient safety [11]; similarly, 3D-printed kidney models can enhance patients’ understanding of their pathology and anatomy, as well as their interaction with healthcare professionals [12]. However, printing costs remain a significant barrier to their widespread use [12,13]. The literature comparing personalized 3D-printed tumoral kidney models to generic tumoral kidney models is sparse, and no randomized studies exist.

## MATERIALS AND METHODS

### Aim

The aim of this study is to investigate the benefits of using a personalized 3D-printed model of the tumoral kidney versus a generic 3D-printed tumoral kidney model as an educational tool throughout the care pathway, on patient’s experience and their interactions with caregivers

### Trial design

We are conducting a prospective, unblinded, randomized trial with two parallel groups (1:1 ratio). Participants will be assigned to either use of a personalized 3D-printed model or a generic 3D-printed model as information and communication support throughout their care journey. The study design is presented in figure 1.

**Figure 1.**
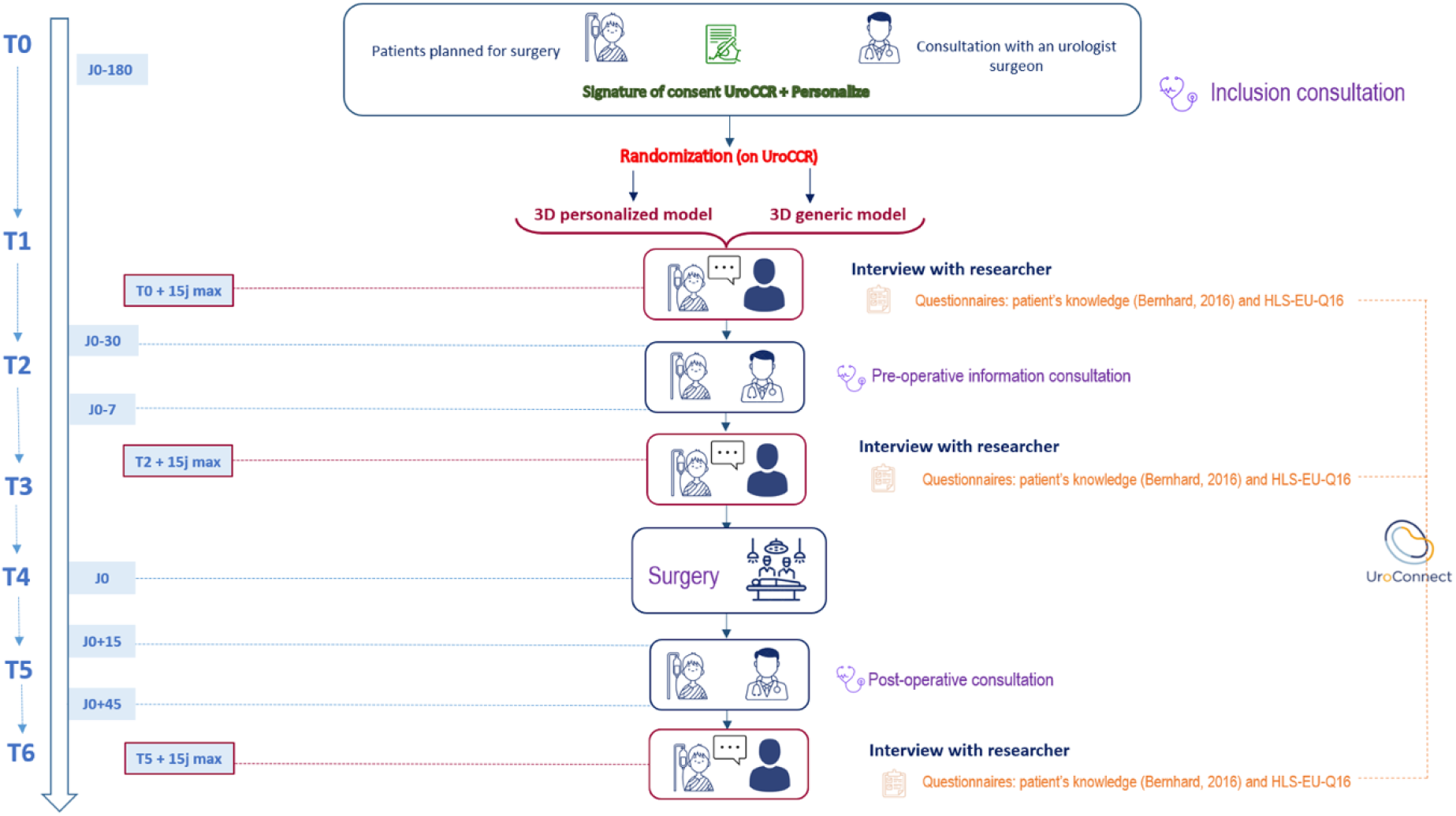
Study design

### Study population

Patients enrollment will take place during standard diagnostic consultations for kidney tumor prior to being scheduled for RAPN at a high-volume academic hospital. Recruitment started in May 2024, inclusion and exclusion criteria are outlined in table 1. Eligible participants will include patients aged 18 or older who are scheduled for RAPN for a unilateral renal tumor, or first surgery for bilateral tumors, and who are affiliated to the French social security system. Patients will be excluded if they are contraindicated to a four-phase preoperative CT scan required to produce the 3D model, have a solitary kidney, a history of other cancers, or metastatic disease at inclusion, are under trusteeship, curatorship, or legal guardianship, or if they refuse to give consent in either the UroCCR project or this trial. All patients and surgical data will be stored in the database of the French research network on kidney cancer UroCCR (NCT03293563). This prospectively maintained database is IRB-approved and obtained the CNIL authorization number DR-2013-206 and EDS authorization DT-2024-027. It has been labeled by the French National Cancer Institute (INCa), the French High Authority of Health (HAS), the French Scientific Interest Group on Biology, Health and Agronomy (GIS-IBiSA) and the French National Research Agency (ANR).

**Table 1.** Inclusion and exclusion criteria *R3DP-P: Rein 3D – Personalize*

| Inclusion criteria | Exclusion criteria |
| --- | --- |
| Male and female aged of 18 and over | Language difficulties in French |
| Scheduled for a first robotic-assisted partial nephrectomy for a unilateral renal tumor, or first surgery for a bilateral tumor | Contraindication to a four-phase preoperative CT scan required to produce the 3D model |
| Affiliation to or beneficiary of the French social security | Person under trusteeship, curatorship, or legal guardianship |
| Free, informed, and written consent signed by the patient and the investigating physician (at the latest on the day of inclusion and before any examination required by the research) | Refusal of consent or participation in the UroCCR project and the R3DP-P ancillary trial |

**Table 2.** SPIRIT figure, schedule of enrolment, interventions, and assessment. T0 to T5 = timepoints 1 to 5, D0 = day of surgery.

|  |  | STUDY PERIOD |  |  |  |  |  |  |  |
| --- | --- | --- | --- | --- | --- | --- | --- | --- | --- |
|  |  | Enrolment | Allocation | Post-allocation |  |  |  | Close-out |  |
| Timepoint |  | T0<br>(First consultation) | T1<br>(T0 + 1 day) | (T1 + 15 days) | T2<br>(30 to 7 days before D0) | T3<br>(T2 + 15 days) | T4<br>(D0) | T5<br>(T4 + 15 to +45 days) | T6<br>T5+15 days |
| ENROLMENT | Eligibility screen | X |  |  |  |  |  |  |  |
|  | Informed consent | X |  |  |  |  |  |  |  |
|  | Allocation |  | X |  |  |  |  |  |  |
| INTERVENTION | Modeling virtual 3D-model (according to randomized arm) |  |  | X |  |  |  |  |  |
|  | Printing 3D-model (according to randomized arm) |  |  | X |  |  |  |  |  |
|  | Preoperative information according to randomized arm |  |  |  | X |  |  |  |  |
|  | Surgery |  |  |  |  |  | X |  |  |
|  | Postoperative consultation |  |  |  |  |  |  | X |  |
| ASSESSMENTS | HLSEU-Q16 questionnaire (health literacy) |  | X |  |  |  |  |  | X |
|  | Questionnaire (knowledge of disease and planned surgery) |  | X |  |  | X |  |  | X |
|  | Questionnaire of satisfaction |  |  |  |  |  |  |  | X |
|  | Semi-directive interview |  | X |  |  | X |  |  | X |

Healthcare professionals involved in patient care will be invited to participate in the study. Participants will include junior and senior urologists, residents, nurses from the urology ward, operating theatre nurses, auxiliary nurses, nurse anesthetists and anesthesiologists. We will ensure diversity in terms of profession and seniority to better represent professional heterogeneity and diversity of experience. An initial interview will take place before the introduction of 3D models in the department, and a second interview will be conducted one year after the study begins.

### Study setting

In this study, patients scheduled for RAPN will be randomized into two groups: one informed using a personalized 3D-printed model of their tumoral kidney and the other a generic 3D-printed model. Consecutive patients who meet the eligibility criteria during the inclusion period will be offered to participate in the study. All participants and health professionals will provide written informed consent prior to their inclusion in both UroCCR and in the Rein 3D print - Personalize (R3DP-P) study. After inclusion, patients’ data will be anonymized and collected in the UroCCR database. Source data from observations (notebook) and interviews (using digital audio recording) will be transcribed, entered, and coded exclusively by the HSS (humanities and social sciences) research team. The transcribed data will be documented directly into the Nvivo Pro 13® qualitative analysis software, and subsequently analyzed using Nvivo Pro 13®.

### Randomization and blinding

Randomization between the two groups is balanced 1:1, without stratification. The randomization list is established by a statistician at Bordeaux University Hospital’s Clinical Epidemilogy Unit before the onset of the study and is implemented in the electronic UroCCR platform. Randomization will be carried out on the UroCCR platform, immediately following the inclusion visit. Upon confirming a patient’s eligibility, the investigator accesses the UroCCR website using secure credentials. The platform then instantly provides the investigator with a unique participant identifier and the result of the randomization process. Although the study is designed as unblinded, patients will remain unaware of their assigned randomization group until their designated follow-up visit.

### Procedure

Each patient will be randomly assigned to one of the two parallel groups (1:1 ratio) at inclusion, corresponding to the information mediation tool used throughout the care pathway. All patients will undergo a high quality, four-phase preoperative abdominal CT scan.

- Personalized 3D-printed model group: CT scans are segmented by a urologist using Synapse 3D® software (Fujifilm) to generate comprehensive 3D models of the tumoral kidney, its vascularization and excretory tract. The models are then printed using high-resolution, multi-material 3D printing technology (Stratasys J750® 3D printer) (Figure 2). The personalized three-dimensional printed physical model of the patient’s tumoral kidney will be used as an educational tool.
- Generic 3D-printed model group: a three-dimensional printed physical model of a tumoral kidney selected from a set of eight models that best matches the patient’s condition will be used as an educational tool. These models were created from the CT-scans of previously operated patients and were selected to represent the full spectrum of kidney cancer presentations. Figure 3 shows the eight kidney models and corresponding RENAL nephrometry scores and TNM staging [14]. Three of the models represented anterior tumors of low to intermediate complexity (Renal scores *5a, 8a and 9a, all cT1a)*, two other models were upper and lower posterior tumors of low to intermediate complexity (Renal scores *8p and 5p, cT1b and cT1a*), hilar endophytic and exophytic (Renal scores *10xh and 11xh, cT1b and cT1a)* and one large inferior tumor (Renal scores *10x, cT2b)*.

**Figure 2.**
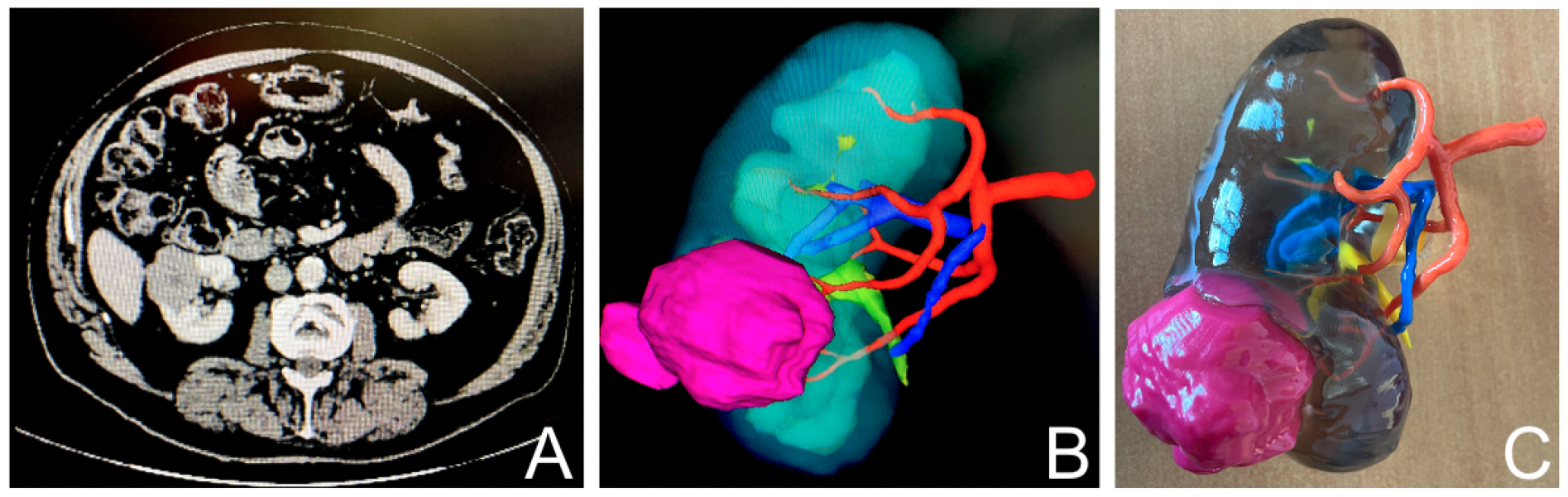
3D printed model workflow: a/CT-scan,b/ 3D model rendering, c/3D printing

**Figure 3.**
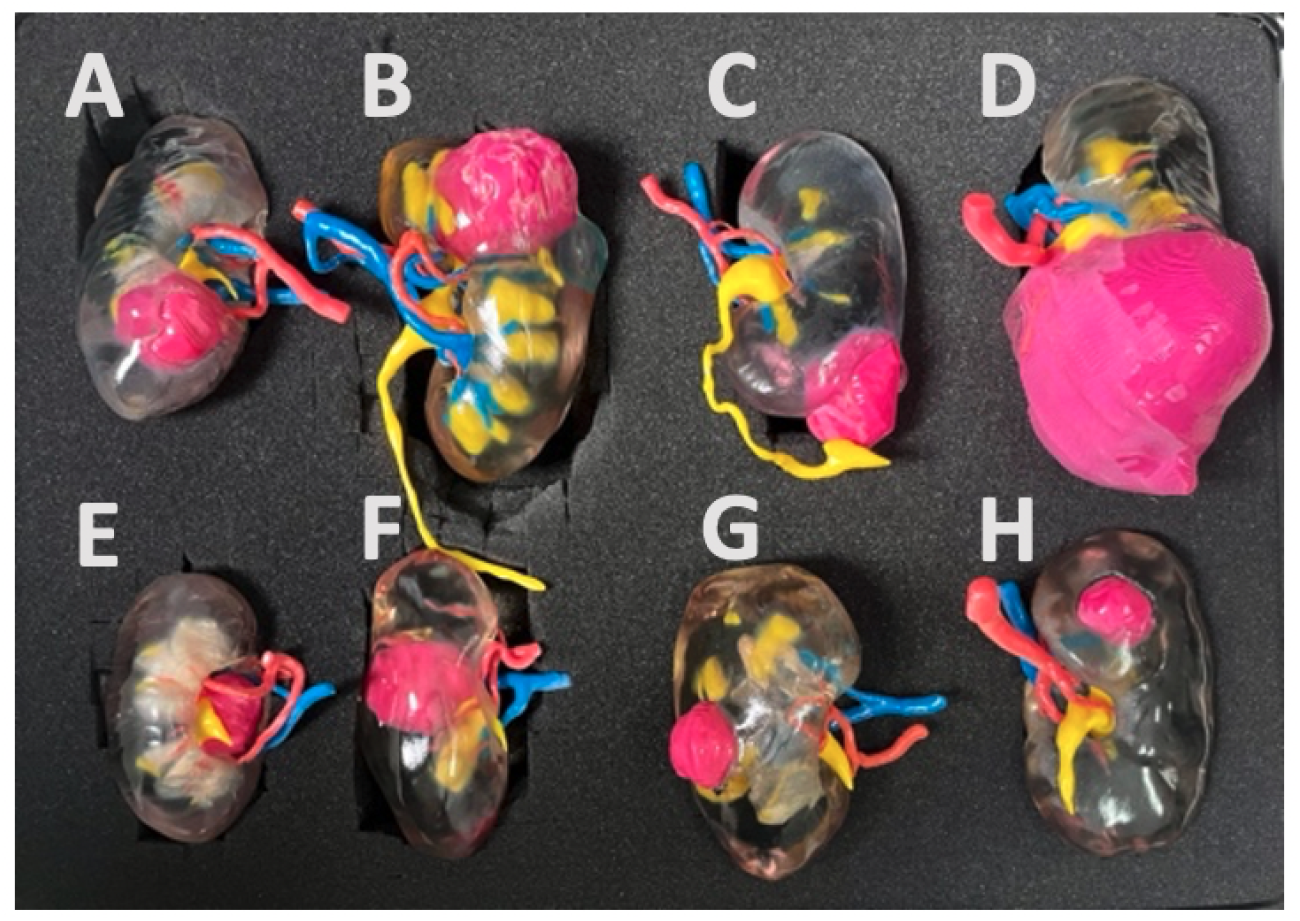
Generic kidney models set (Renal scores [14] according to each kidney : *A*.*8a B*.*9a C*.*5a D*.*10x E*.*10xh F*.*11xh G*.*8p H*.*5p)*

A surgical consultation using the 3D-model from the patient’s allocated group will take place between 30 and 7 days before the surgery. This consultation serves to inform the patient about their medical situation and the planned operation, using the 3D model as an educational tool. It allows patients to ask any questions, express doubts or clarify misunderstandings they may have. Importantly, the explanations provided during this consultation are not standardized and follow routine clinical practice, allowing for natural dialogue and variation across interactions. The 3D model then accompanies the patient throughout their care pathway to serve as a tool for communication and education.

### Outcomes measures

All patients will complete questionnaires assessing their health literacy, knowledge of kidney-tumor anatomy, and satisfaction with the use of 3D-printed kidney models as educational tools. Patients’ experience and their interactions with caregivers will be analyzed during semi-structured interviews conducted at three different times: after the first consultation with the surgeon and before the preoperative education consultation, between this second consultation and the surgery, and after the post-operative consultation. The impact of these 3D models on healthcare professionals and the changes induced in their practice will be assessed through two interviews: one interview before the start of the study, i.e., the introduction of 3D models into the care pathways, and a second interview one year after the introduction of the models.

#### Primary outcomes

Qualitative evaluation of patients’ experience using a personalized 3D-printed kidney model versus a generic 3D-printed kidney model as an education tool throughout the care pathway, focusing on patient experiences and their interactions with healthcare professionals before and after partial nephrectomy. These experiences will be analyzed in conjunction with clinical and demographic data from the UroCCR database.

#### Secondary outcomes

- Detailed description of interactions between caregivers and patients, including the context, environment, practice, vocabulary, and discourse content used throughout the care process.
- Comparison of speech and terminology differences when using the different models during interactions between patients, caregivers, and patients’ families, through systematic analysis of interviews.
- Investigation of the 3D-printed kidney model’s use from the initial consultation to the post-operative visit, documenting its role throughout the entire care process.
- Comparison of patient’s understanding of anatomy and surgical strategy based on the type of model used (personalized vs generic) before and after the surgery (Supplementary material 1).
- Comparison of health literacy levels between the two groups at various times, analyzing changes in the average score on the Health literacy Survey Questionnaire (HLSEU-Q16: European Health Literacy Survey Questionnaire [15], French version [16]) (Supplementary material 2).
- Evaluation of Healthcare professionals’ adoption of the 3D tool and perception of patient management following the integration of 3D models, by qualitatively assessing changes in practices throughout the care process.

All questionnaires will be distributed using the UroConnect® application (Resilience) [17], a telemonitoring tool routinely used in our nurse-led pathways (enhanced rehabilitation after surgery and ambulatory) or in paper format for patients without internet access. The application will notify participants at designated times to complete the appropriate questionnaire.

### Anthropology-driven approach

We will employ a comprehensive health anthropology-driven [18] approach to examine the information and educational interactions between healthcare professionals and patients. The following interactions will be investigated by two educational experts:

- The way caregivers provide information to patients, exploring patient experiences, and assessing health education and literacy [19].
- The informational context that occurs within the patient care pathway, describing the process of knowledge dissemination and construction of various participants, and examining the adaptation work performed by professionals during interactions with patients.

Additionally, a quantitative approach will complement our comprehensive and didactic exploration, allowing us to question the impact of integrating the model on patients’ understanding of their pathology and their level of health literacy. This will be evaluated using the validated HLSEU-Q16 literacy questionnaire (Supplementary material 2) and the pathology understanding questionnaire previously used by Bernhard et al. [1] (Supplementary material 1). This study will employ a mixed-method design, combining qualitative and quantitative elements. It is a single-center study based on observations (interaction situations), interviews (with professionals and patients), and questionnaires (completed by patients).

### Pre- and post-operative follow-up

Study participation begins with an inclusion consultation (T0), aligning with standard management practices as depicted in the timeline in figure 1. Following this consultation, patients are randomized into one of two groups, allowing the surgical team to create a personalized 3D printed model of patients’ tumoral kidney for those included in the personalized group.

Subsequently, an interview with the HSS researcher is conducted either at the hospital or at home (T1), according to the patient’s preferences, within 15 days after the inclusion visit. This face-to-face meeting explores the patient’s experience from the initial onset of symptoms to seeking urological care. It also gathers socio-demographic data and insights into the patient’s interactions with family and peers regarding the disease. During this visit, patients will complete questionnaires via the UroConnect platform, focusing on HLSEU-Q16 literacy, and their understanding of renal anatomy and surgical issues considerations.

Fifteen days after the “3D model introduction” consultation (T2), patients will participate in another interview (T3), either at the hospital or at home. This session gathers insights about their experience since the previous interview and their interactions with healthcare professionals, family, and friends related to the disease. At this point, both questionnaires will be completed again. The surgery will then be performed, 7 to 30 days after the T2 consultation, without altering the surgeon’s standard practice.

The collection of research data will conclude within 15 days after the one-month post-operative consultation. A final interview will be conducted either at home or the hospital, depending on the patient’s preference. This meeting will explore the patient’s experience since the prior interview and their communications with relatives about the disease. Participants will also complete one last time the questionnaires via the UroConnect platform, addressing HLSEU-Q16 literacy, their understanding of renal anatomy and surgical issues, and their overall satisfaction.

### Statistical analysis

#### Sample size

To integrate both qualitative (observations, interviews) and quantitative (questionnaires) methodologies, the study was designed to include 60 patients, divided into two randomized groups of 30 each. Should any patients be lost to follow-up, they will be excluded from the study and replaced, with a cap on total inclusion at 68 participants. Furthermore, interviews will be conducted with 30 healthcare professionals who meet the inclusion criteria.

#### Data analysis

A mixed-methods analysis will be employed using a triangulation approach. Sociodemographic data and clinical characteristics of the patients will be described in terms of frequencies for the qualitative variables and means with standard deviations for the quantitative variables and compared between the two groups. Data from patients’ questionnaires will undergo descriptive analysis, detailing the score distribution for the HLS-EU16 literacy questionnaire at baseline and study conclusion, as well as satisfaction with the 3D models utilization.

The analysis of patients’ experiences will be informed by content from cross-sectional interviews at each stage, alongside a longitudinal study of their progression. Changes in understanding of the disease and surgical process will be examined through interviews and questionnaires content, with comparative analyses between the two groups. Additionally, content from healthcare providers interviews will be analyzed.

### Trial status

The protocol number is ID-RCB 2023-A00146-39, version no. 1 from 27/10/2023. Recruitment began in May 2024. Inclusion deadline is scheduled September 2025. Trial is registered with the reference NCT06035211 [ClinicalTrials.gov] [registered on 5 September 2023]. UroCCR network is registered with the reference NCT03293563.

## Discussion

We hypothesize that both 3D tumoral kidney models will serve as a crucial educational tool during the patient’s interactions with relatives and healthcare professionals: they are intended to empower the patient by integrating it as a part of their healthcare process. The personalized 3D printed model may facilitate tailored discussions about the patient’s specific condition while enhancing the understanding of their illness. Thus, the introduction of these 3D models could have an impact on the relationship that professionals have with patients.

We furthermore assume that patients’ ability to visually and physically interact with the 3D representation of “their” kidney will significantly enhance their comprehension of their pathology throughout their care pathway. This hypothesis is driven by the belief that tangible, personalized representations can aid in demystifying complex medical conditions for patients.

## Data Availability

Data will be coded and hosted on dedicated servers with secure access managed by the CREDIM (medical informatics research and development center – Bordeaux University). Only the database managers, the research team and the auditors will have directly access to the data. Investigators will make data available to persons with access to these documents, in accordance with the legislative and regulatory requirements in effect.

## Abbreviations

RAPN: (Robotic Assisted Partial Nephrectomy)
R3DP-P: (Rein 3D Print - Personalize)
HSS: (Humanities and Social Sciences)
3D: (Three Dimension)
T0 to T6: (consultation Timepoints)

